# A mathematical model for repetitive behaviors of Covid-19

**DOI:** 10.1101/2021.11.08.21266099

**Authors:** Yoshiro Nishimoto, Kenichi Inoue

## Abstract

Covid-19 pandemic waves have been hitting us again and again in the past couple years in many countries, while the reason why they come in such repetitive manners remains unexplained, which have brought us with lingering anxieties and economic stagnations.

We proposed a mathematical model to describe the mechanism of the repetitive appearance of the number of new cases based upon the *SIQR* model in which *Q* (quarantined infectors) were distinguished from *I* (un-quarantined ones). The repetitive behavior of the pandemic was simulated by an activator-inhibitor system around a fixed point in a phase space as a kind of self-organized oscillations. Periods between each wave were confirmed to be approximately similar. Repetitive behaviors were also observed in actual Covid-19 data.

Practical policies and actions were discussed on the ways to effectively control the repetition of pandemic, and proactive PCR test especially after the peak-out stage is highly recommended.

## 1. Introduction

Covid-19 pandemic waves have been hitting us again and again in the past couple years in many countries. Japan was no exception and hit by five waves since April 2020 up to October 2021. Every time the infection clusters show up, experts issue a warning of rapid spread, the government declare a state of emergency and, consequently people are requested to stay. The pandemic wave, sooner or later, settles down and gets started again while the reason for such repetitive behavior of Covid-19 infection has not been nicely explained by the experts of infectious disease.

The number of new cases in countries (Fig.1)^1)^ show that the repetition seems to have little correlation with each government policy such as the state of emergency, city-lockdown or the progress of vaccination rate. Therefore we considered that the repetitive behavior might come from more fundamental mechanisms.

**Fig.1.**
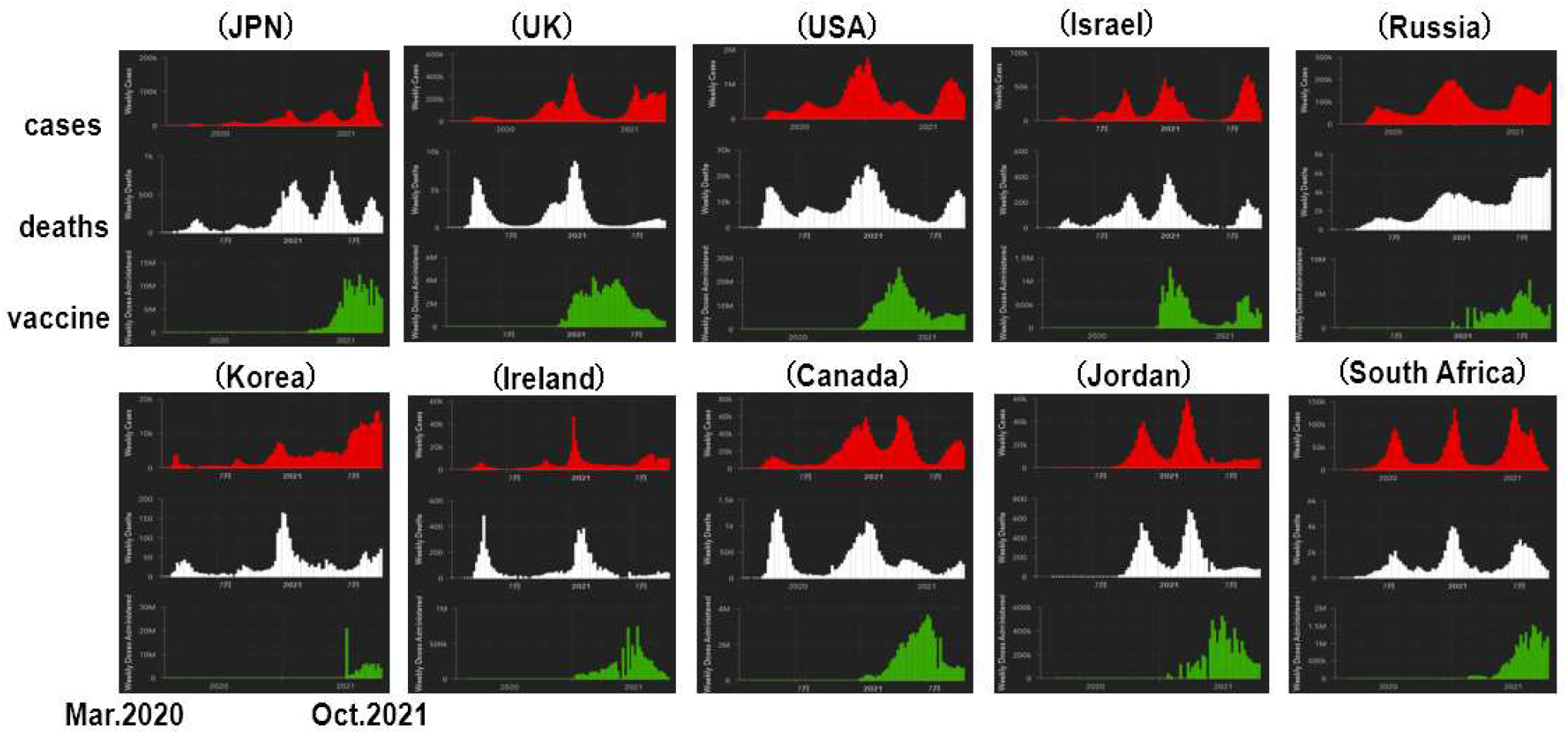
28-day cases, deaths and vaccine doses in countries

We propose a mathematical model to describe the mechanism based upon an idea that the infection is controlled by both infectivity and quarantine rate using the *SIQR* model in which *Q* (quarantined infectors) are distinguished from *I* (un-quarantined ones).

## 2. Mathematical model

The *SIQR* model^2)^ at an earlier pandemic stage far from herd immunity (*S*/*S*(0)∼ 1) can be written as follows;

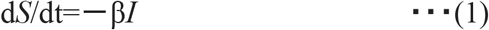

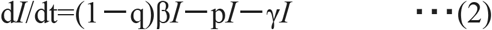

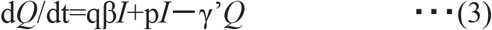

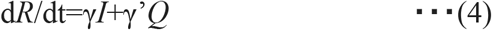

where *S*:a stock of the susceptible, *I*:a stock of the un-quarantined infectors, *Q*:a stock of the quarantined ones and *R*:a stock of the recovered or deaths, β: an infectivity, q and p:quarantine rates and γ:a recovery rate. Eqs.(1)+(2)+(3)+(4) makes d(*S*+*I*+*Q*+*R*)/dt=0 and *S*+*I*+*Q*+*R*=const. Eq.(1) means that β*I* stands for daily generated infectors including both the quarantined and un-quarantined. Eq.(2) means *I* can be approximately expressed by exp(αt),where α=(1− q)β− p− γ…(5). Eq.(3) means that (qβ*I*+p*I*) stands for newly quarantined infectors, of which qβI can be regarded as a real-time index n corresponding to daily generated, PCR tested positive and quarantined infectors. Hence,

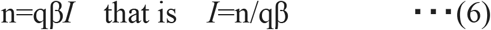

By substituting Eq.(6) into Eq.(2), we have

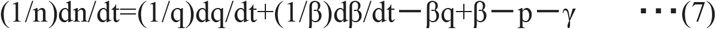

Eq.(7) shows how index n moves interactively with q and β in a phase space q-β-n. Since (1/kx)d(kx)/dt=(1/x)dx/dt, q, β and n are scalable and easy to handle in a mathematical model. In addition, since both q and p are quarantine rates with different populations, they can be regarded as the similar indexes. If p is replaced by q, Eq.(7) can be written as follows;

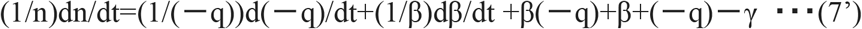

which implies that β and q have an anti-symmetrical effect on index n.

## 3. Simulations by the mathematical model

### (1) Simulation on β-n plane

Firstly under the condition that q is constant in Fig.2, Eq.(7) gives

**Fig.2.**
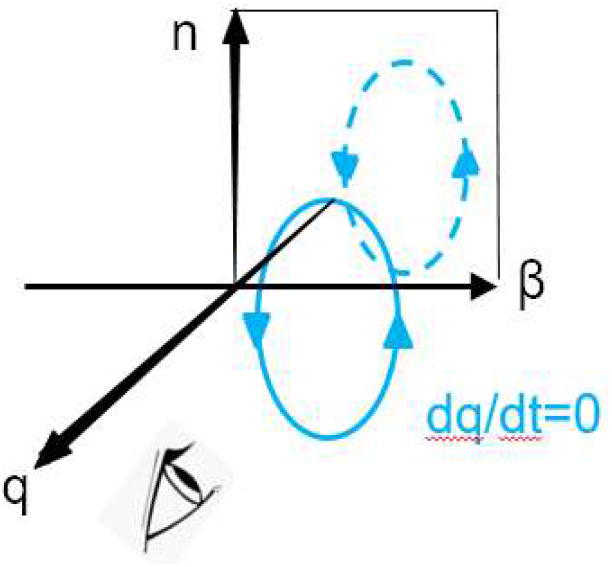
Projection onto β-n plane

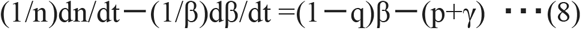

which is non-linear with respect to β and n, and cannot be generally solved. We adopt one of sufficient conditions with undetermined coefficients a,b,c,d,e and f.

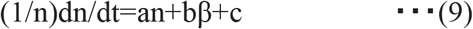

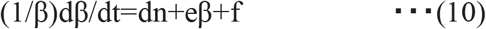

Comparison between Eq.(9)-(10) and (8) gives the following velocity vector (dβ/dt,dn/dt).

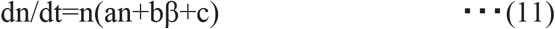

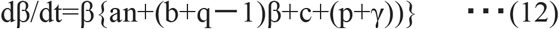

Eq.(11),(12) form an activator-inhibitor system in a limited region of a<0 and b>0 and can simulate an repetitive behavior on the β-n phase plane. Here the so-called ‘nullcline’ is defined by dβ/dt=0 and/or dn/dt=0 ^3)^.

When epidemic parameters q=0.22, p+γ=0.2 and undetermined coefficients a=− 1, b=1, c= − 0.1 for example, the equations and nullclines are as follows;

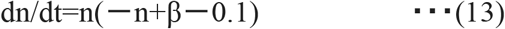

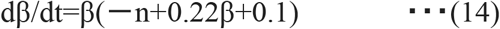

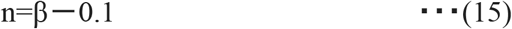

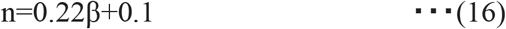

Anti-clockwise rotational field around the fixed point (0.26,0.16) is observed as shown in Fig.3. A locus generated by successive velocity vectors from a starting point (0.7,0.15) is overwritten. A slowly shrinking spiral around the fixed point is well simulated.

**Fig.3.**
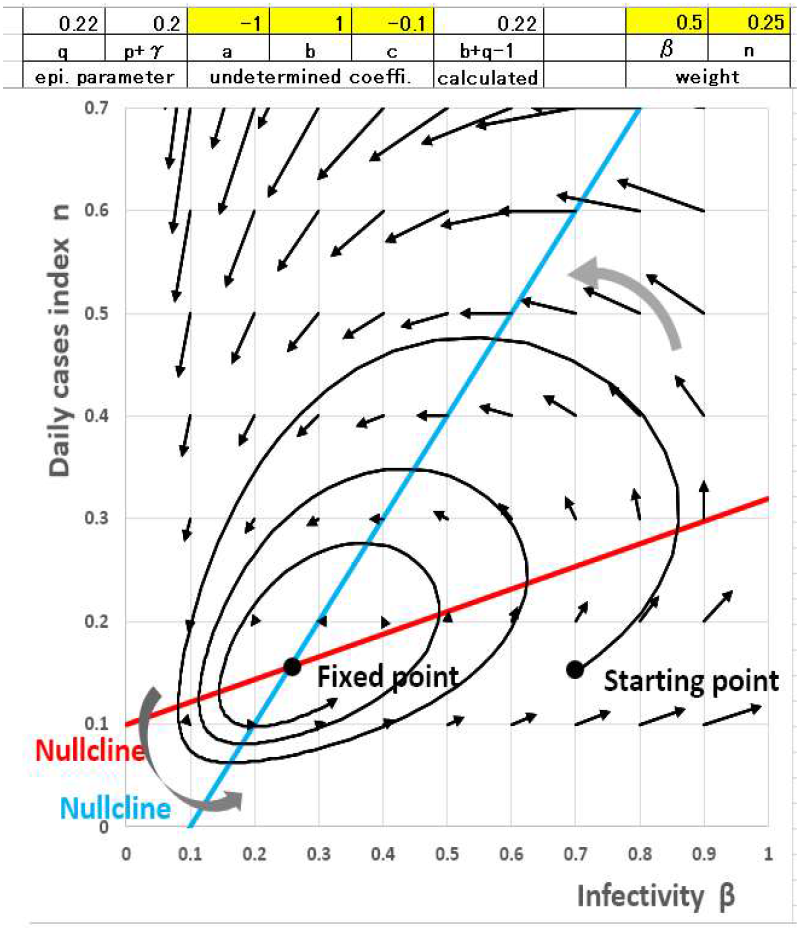
Repetitive behavior on β-n phase plane

The profile of the infectivity β and daily cases index n along the time axis are described in Fig.4, where β is in advance of index n. While remembering the exponential coefficient α=(1− q)β− γ− p…(5), it can be mentioned in terms of β that;

**Fig.4.**
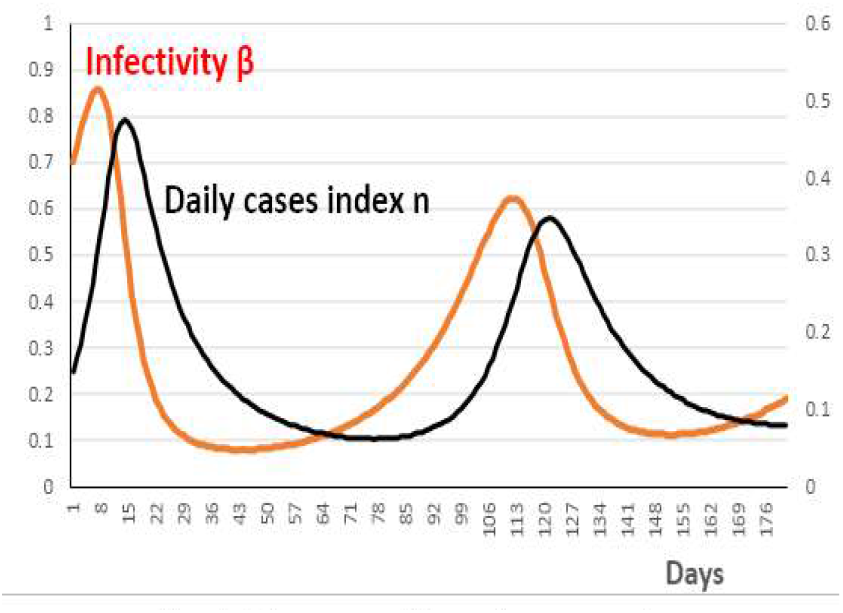
Change of β and n over days

1. At the beginning of a pandemic wave, β is generally so high that positive α makes index n increase exponentially.
2. After then, β is rapidly reduced owing to such as the people’s stay-at-home restriction and local saturations of infection itself.
3. When β is reduced, negative α makes index n peaked out and decreased.
4. We hope that the locus will stay at the region close to the origin of the phase plane. However it will step over nullclines onto the region of the starting point. The reason of this returning behavior is currently unknown, but infectivity β tends to grow bigger again with certain reasons.

### (2) Simulation on q-n plane

On the other hand, under the condition that β is constant in Fig.5, Eq.(7) gives

**Fig.5.**
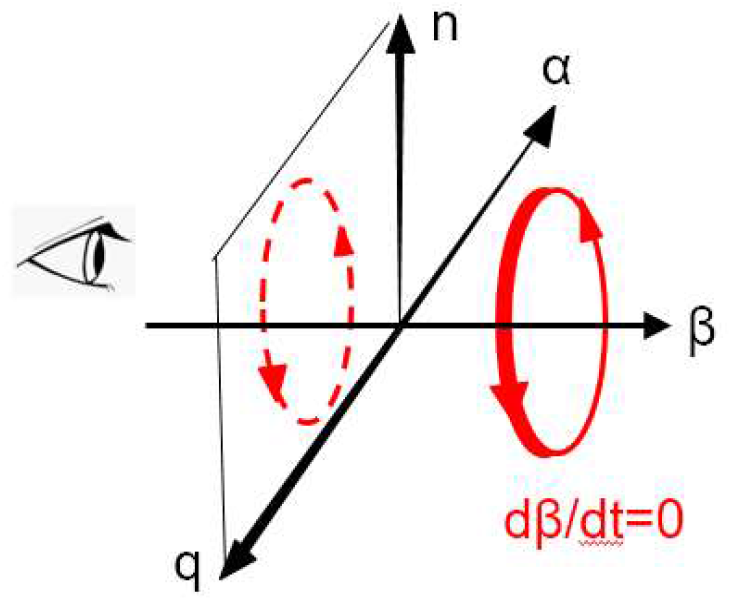
Projection onto q-n plane

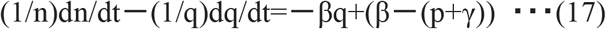

The following velocity vector (dq/dt,dn/dt) is introduced on a q-n plane in the same way as on the β-n plane.

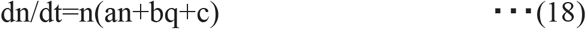

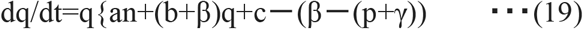

Eq.(18),(19) form an activator-inhibitor system in a limited region of when a>0 and b<0.

A clockwise rotational field around the fixed point (0.38,0.38) is observed as shown in Fig.6. A locus generated by successive velocity vectors from a starting point (0.2,0.4) is overwritten. A slowly shrinking spiral around the fixed point is well simulated.

**Fig.6.**
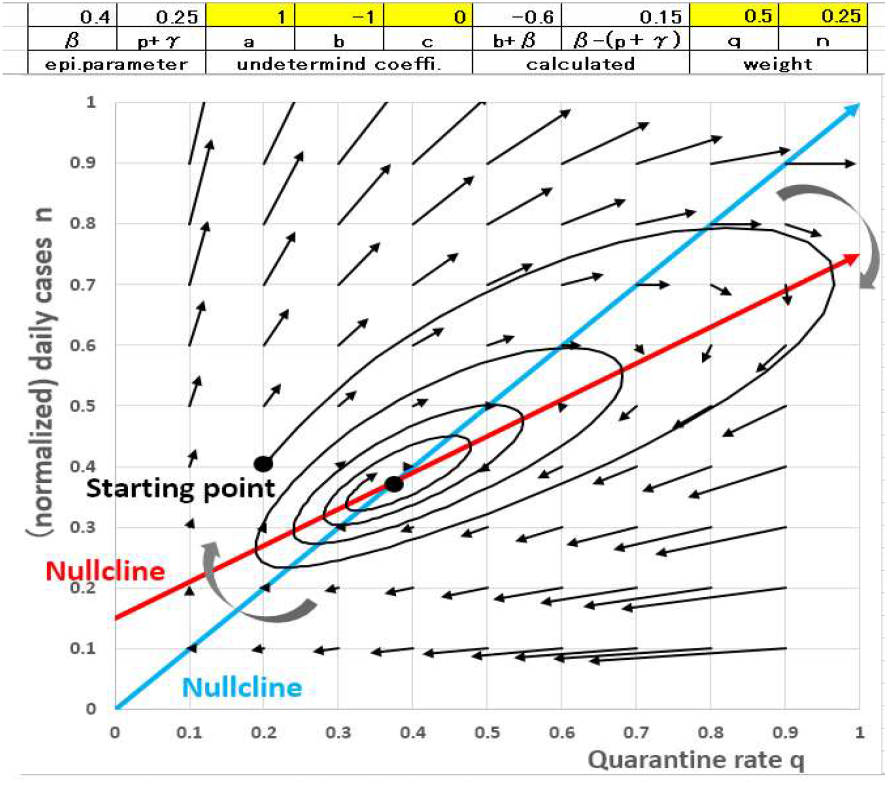
Repetitive behavior on q-n phase plane

The repetitive profile of the quarantine rate q and daily cases index n along the time axis are described in Fig.7, where q is lagging behind n.

**Fig.7.**
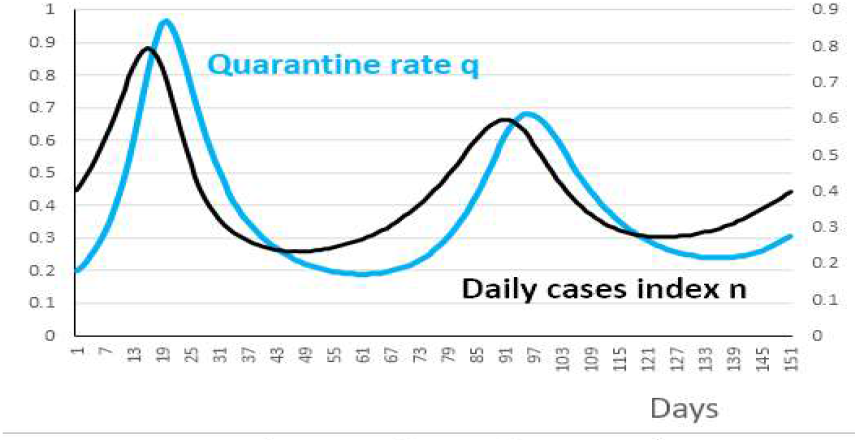
Change of q and n over days

1. Whenever index n increases and close contacting persons come out one after another, PCR tests are performed actively and the quarantine rate q is improved consequently.
2. When q is improved, n rather decrease because quarantined and isolated people do not spread viruses any more.
3. When n decreases, people get more or less relieved and PCR tests are naturally reduced.
4. The reason why the locus steps over the nullclines is unknown. However few PCR tests fail to reduce the quarantine rate q. We have to point out that un-quarantined “hidden infectors” might cause the upcoming wave.

### (3) Integration of β-n and q-n planes

Comparison between Fig.3 and Fig.6 shows that the epidemic loci on the β-n and q-n phase planes have opposite directions as integrated in Fig.8. This is corresponding to the anti-symmetrical properties of β and q against n in Eq.(7’).

The repetitive behaviors derived from the non-linear differential equation (7) can be regarded as a kind of ‘self-organized oscillation’^4)^ in the non-equilibrium pandemic state far from the herd immunity.

**Fig.8.**
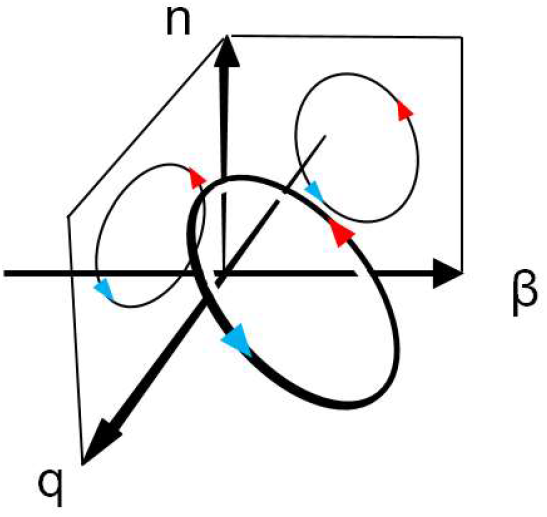
Pandemic locus in q-β-n space

## 4. Discussions

Although the proposed model is one of possible mathematical solutions of the *SIQR* equation, there is no necessity for the actual Covid-19 behaviors to choose the mathematical model. Therefore we have to verify the correspondence between actual data and the model

### (1) Comparison of period between actual and model

The Covid-19 five waves in Japan have been repeated roughly periodically with peak-to-peak periods of 125 ±22 days in spite of different peak heights as shown in Fig.9. What is the period T between neighboring peaks of the repetitive loci in the model? T can be calculated using the following path integral.

**Fig.9.**
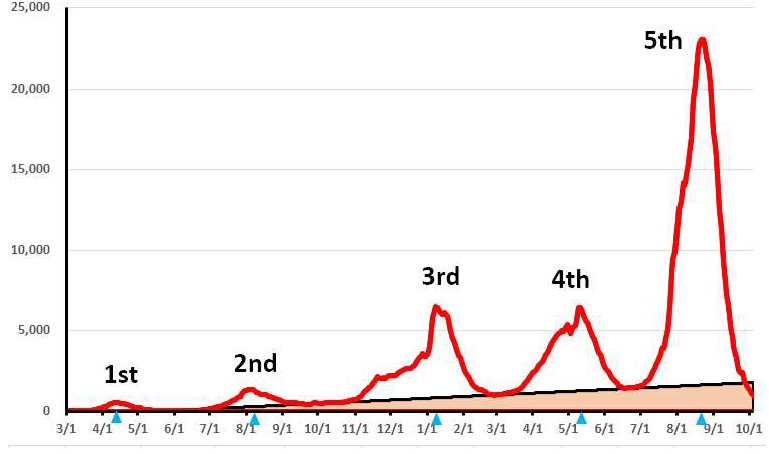
Covid-19 cyclic waves in Japan

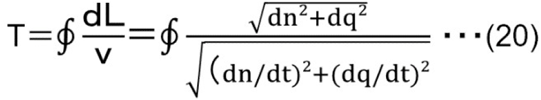

where (dq/dt,dn/dt) is the velocity vector on the q-n plane. Although the spiral locus includes large, middle and small turns described in Fig.10, each period T1,T2, and T3 is almost the same as shown in Fig.11. It was also found that it took a considerable number of days for the locus to step over the lower side of the nullclines.

**Fig.10.**
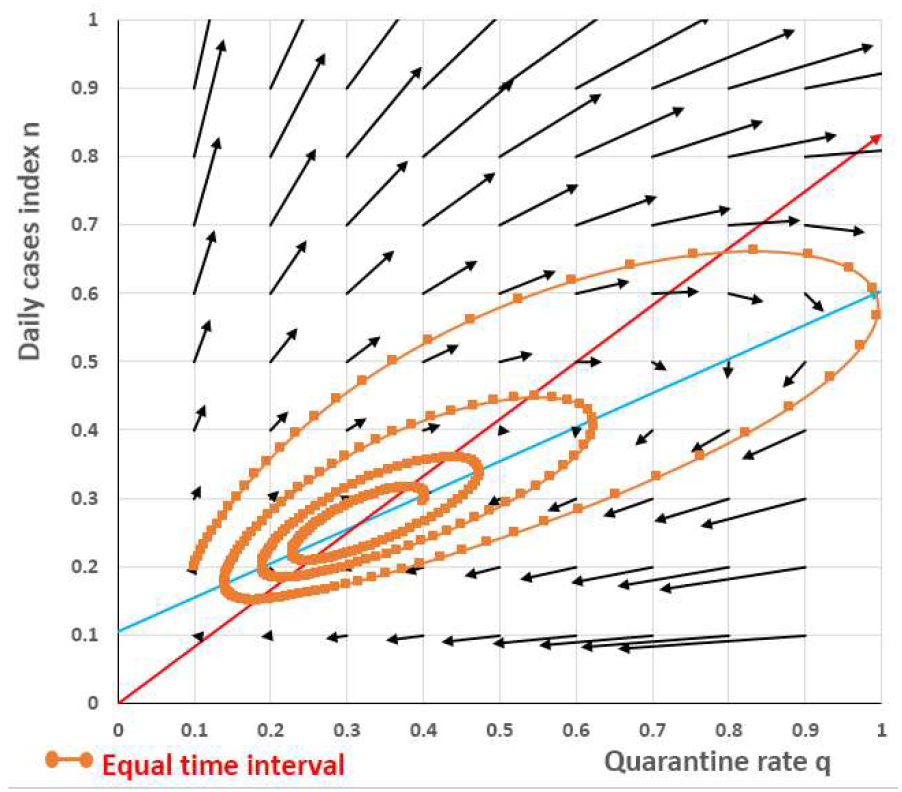
Time analysis by path integral on q-n plane

**Fig.11.**
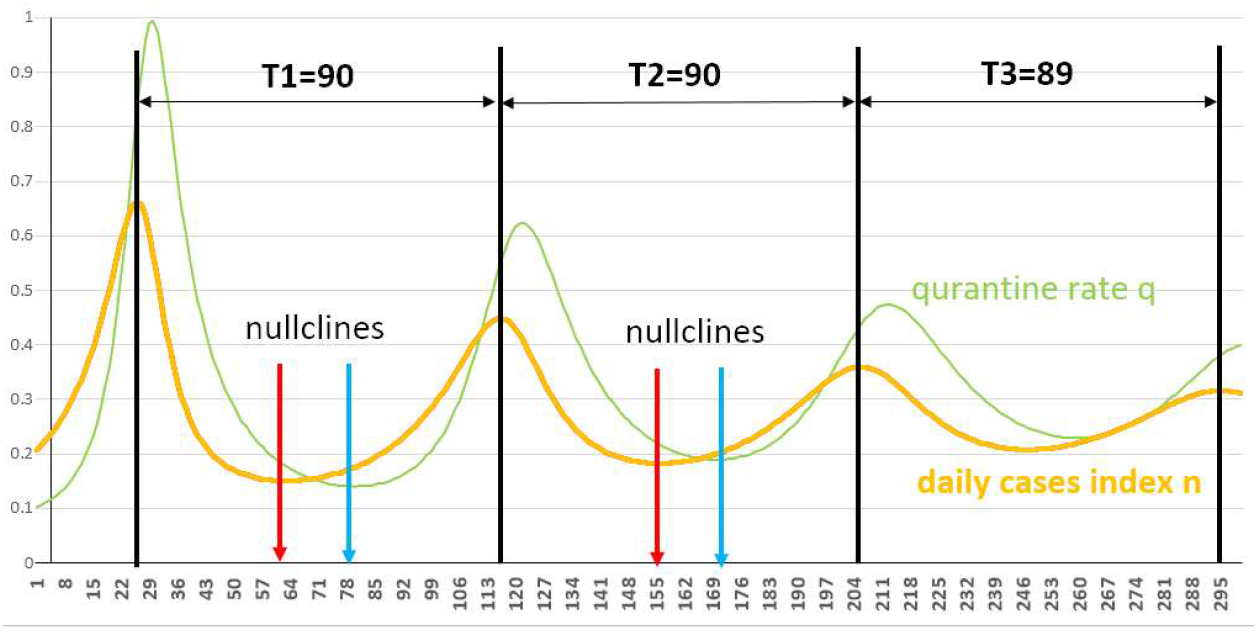
Evaluation of period between peaks

### (2) Repetitive behaviors with public data

It is hard to find quarantine rate q directly by solving the non-linear differential equation (7). We use the number of PCR tests (hereafter represented by ‘PCR’) in place of q since un-quarantined infectors *I* are roughly proportional to the positive rate in PCR tests (n/PCR) and from Eq.(5), q=n/β*I*∝ PCR. Repetitive patterns can be observed for the 3^rd^, 4^th^ and 5^th^ waves in Japan in Fig.12, and 3^rd^ waves in UK and USA and 2^nd^ wave in South Africa in Fig.13, respectively on PCR-n planes.^5)^

**Fig.12.**
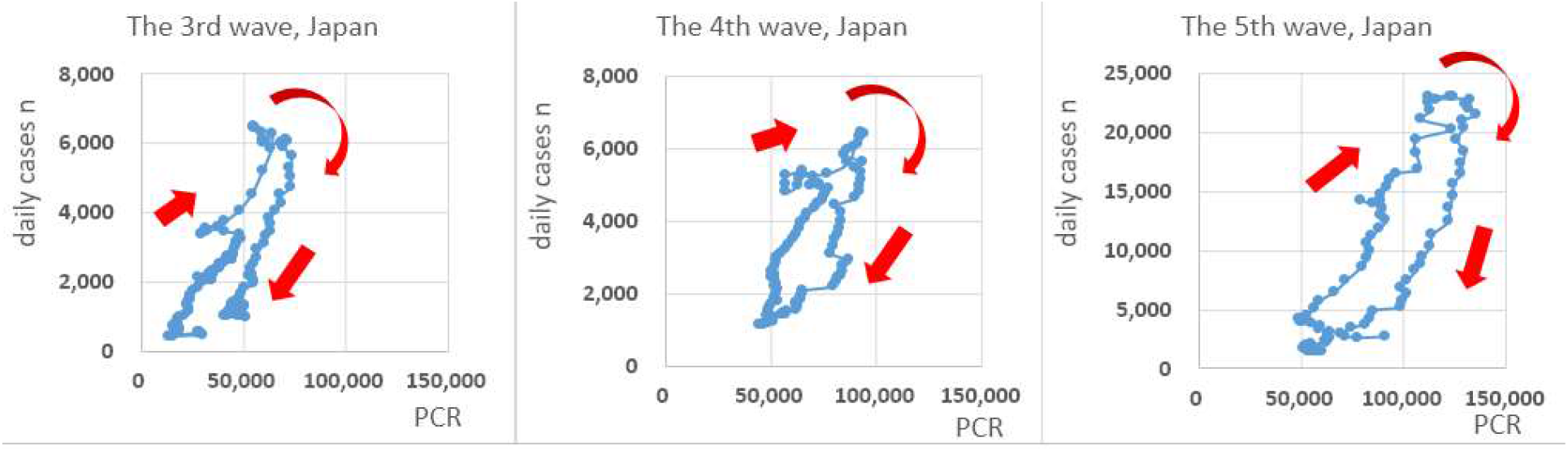
Repetitive patterns on PCR-n plane observed for waves in Japan

**Fig.13.**
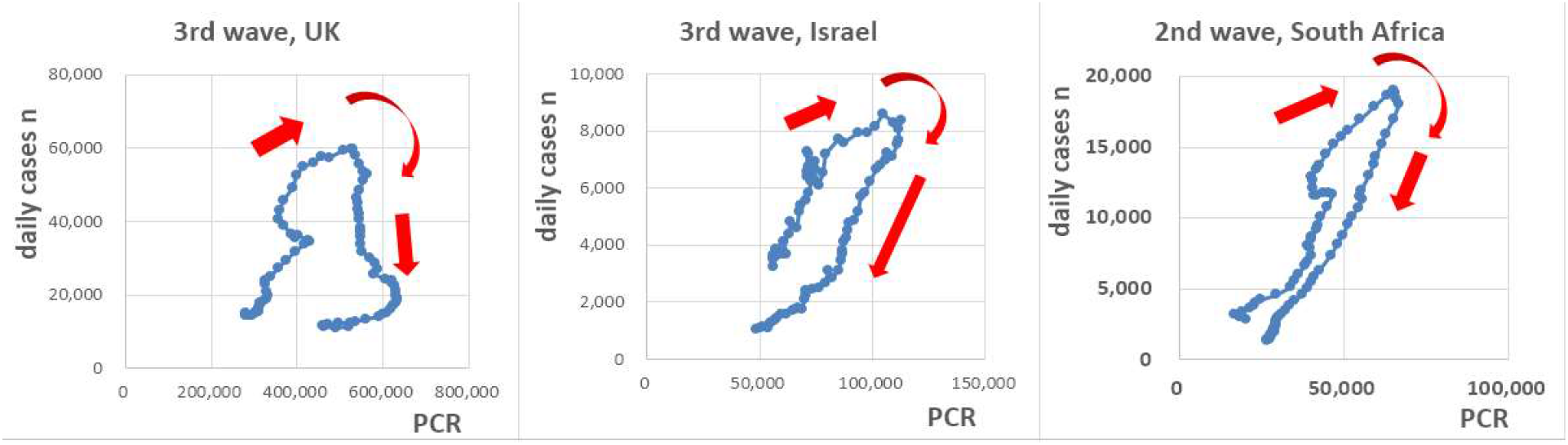
Repetitive patterns on PCR-n plane for waves in UK, Israel and South Africa

### (3) How to avoid or mitigate next wave to come

Overviewing the q-β-n space in Fig.8 (re-posted below) suggests how a Covid-19 wave gets started, developing, peaked-out and settled in terms of β and q schematically in Fig.14. β and q will complementarily activate and inhibit the pandemic wave. Although the mechanism of the next wave trigger is unknown so far, it is true that both β and q become less intense at the same time when daily cases index n is settled down. Therefore we could hope that we would avoid the next wave by keeping the infectivity β low and enhancing the quarantine rate q.

**Fig.14.**
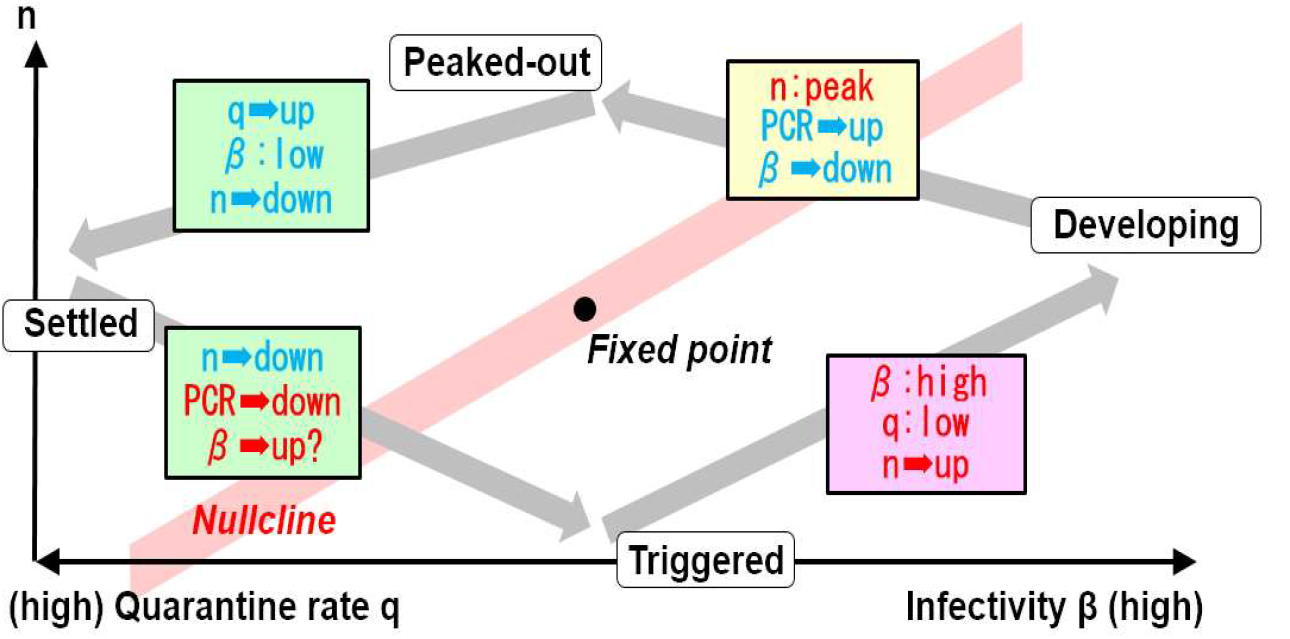
Actual pandemic stages vs. β and q

**Fig.8.**
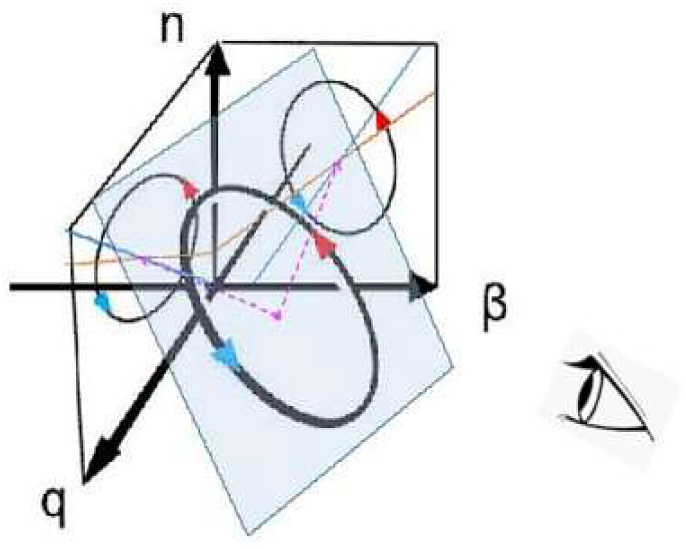
Pandemic locus in q-β-n space (re-poseted)

Reducing of the infectivity β has already been performed through various policies such as face coverings, stay-at-home restrictions, city lockdowns and vaccinations while enhancing the quarantine rate q has not been systematically carried out yet since q is not easy to follow because the “hidden infectors” cannot be correctly estimated.

However we empirically know that proactive PCR test is a good screening method to find “hidden infectors” and enables to quarantine them in a smoother way. For example China has been carrying out a whole city-scaled PCR test which seems successful in suppressing the pandemic spread under the governmental power politics. Since there is no chance of deploying such power politics in Japan, every effort for proactive PCR test is highly recommended for screening and quarantine the “hidden infectors” which should prevent them from becoming a trigger for the next wave. When the wave in certain area seems settled, it is exactly the time to push ahead with such actions.

## 5. Conclusions

A mathematical model based upon *SIQR* model was proposed to understand the mechanism of repetitive appearance of Covid-19 pandemic.

1. A pair of equations forming an activator-inhibitor system were introduced and repetitive behaviors were well simulated in velocity vector fields on the β-n, q-n phase plane and consequently in the q-β-n phase space.
2. Correspondence between Covid-19 actual data and mathematical model data were investigated and, a similarity in the periodicity was confirmed. Repetitive behaviors were also found on the PCR -n plane for pandemic waves in Japan and in some other countries. The model, however, is still on its way to be verified with actual data in other countries across the world.
3. In addition to the government policy of reducing the infectivity β, every effort for proactive PCR test is highly recommended to screen and to find “hidden infectors” in cities and to prevent them from becoming a trigger of the next wave.

## Data Availability

All data produced in the present study are available upon reasonable request to the authors

https://science-research.p-kit.com/page0002.html

## Acknowledgements

The authors thank Prof. T. Odagaki for his instructive advice on his *SIQR* model and Dr. Y.Yamaguchi for his timely suggestion on non-linear pattern dynamics.

